# Cutaneous Microvascular Functional Reserve is Associated with Kidney Function and Histopathologic Injury in CKD: The MAP-CKD Study

**DOI:** 10.64898/2026.04.24.26351712

**Authors:** Armin Ahmadi, Masfiqur Rahaman, Amol Harsh, Jason Yang, Basma Ghanim, Subhasis Dasgupta, Robert N. Weinreb, Tauhidur Rahman, Alfons JHM Houben, Joachim H Ix, Rakesh Malhotra

**Affiliations:** Division of Nephrology-Hypertension, Department of Medicine, University of California San Diego, San Diego, California, USA; Halıcıoğlu Data Science Institute, University of California San Diego, La Jolla, California, USA; Mohamed bin Zayed University of Artificial Intelligence, Abu Dhabi, United Arab Emirates; San Diego Supercomputer Center, University of California San Diego, La Jolla, California, USA; Viterbi Family, Department of Ophthalmology and Shiley Eye Institute, University of California San Diego, San Diego, California, USA; Department of Internal Medicine and Cardiovascular Research Institute Maastricht, Maastricht University Medical Center, Maastricht, The Netherlands; School of Public Health, University of California San Diego, San Diego, California, USA; Nephrology Section, Veterans Affairs San Diego Healthcare System, La Jolla, California, USA

## Abstract

**Background:** Microvascular dysfunction is a key contributor to the development and progression of chronic kidney disease (CKD), yet direct and reproducible assessment of microvascular function in clinical CKD populations remains limited. Laser Doppler flowmetry (LDF) provides a noninvasive, dynamic assessment of skin microvascular blood flow and may serve as a surrogate measure of systemic microvascular health. However, the extent to which LDF-derived measures relate to kidney function, proteinuria, and kidney histopathology in CKD remains unclear.

**Methods:** We assessed cutaneous microvascular function in 150 participants with CKD (estimated glomerular filtration rate [eGFR] <90 mL/min/1.73 m²) using a standardized forearm LDF protocol. Baseline perfusion was recorded at ∼30°C, followed by local heating to 44 °C to induce hyperemia. The percentage change in perfusion unit (PU) was calculated and used to define microvascular functional reserve. Associations between LDF-derived measures with eGFR and urine protein-to-creatinine ratio (uPCR) were assessed using multivariable linear regression adjusted for demographic and clinical covariates. Unsupervised k-means clustering was performed to identify microvascular phenotypes based on resting PU and microvascular function reserve. Associations of LDF measures with glomerulosclerosis (GS) and interstitial fibrosis and tubular atrophy (IFTA) were evaluated in a subset of participants (n = 20) who underwent clinically indicated kidney biopsies.

**Results:** Among 150 CKD participants, the mean (SD) age was 64 (14) years, 46% were female, 38% had diabetes, and 83% had hypertension. The mean eGFR was 42 (21) mL/min/1.73 m² and median uPCR was 0.21 (interquartile range (IQR) 0.11 to 1.20) mg/mg. Higher baseline PU (β = -12; 95% CI, -24 to -1) and reduced percentage change in PU (β = 7; 95% CI, 2 to 13) was associated with lower eGFR, independent of covariates. Baseline PU or percentage change in PU were not associated with uPCR. Unsupervised clustering identified four distinct microvascular phenotypes characterized by graded differences in resting perfusion and microvascular function reserve. Among participants with biopsy data, higher baseline PU and lower percentage change in PU were associated with greater severity of GS and IFTA.

**Conclusion:** In persons with CKD, elevated resting perfusion and impaired microvascular functional reserve were associated with lower eGFR. These findings suggest that LDF-derived measures capture clinically relevant alterations in systemic microvascular function and may serve as a noninvasive biomarker of kidney function and underlying histopathologic injury in CKD.

## Introduction

Kidney biopsy studies have confirmed the microvascular disease as pathophysiologic hallmark of chronic kidney disease (CKD).^1,2^ Emerging evidence indicates that microvascular dysfunction often precedes macrovascular disease, and plays a central role in development and progression of CKD.^2^ These vascular changes are thought to arise from shared pathophysiologic mechanisms, including chronic inflammation, oxidative stress, tissue hypoxia and endothelial injury.^3,4^ Endothelial dysfunction represents one of the earliest and potentially reversible manifestations of microvascular impairement.^5^ Despite its importance, direct and reproducible assessment of microvascular function in clinical settings remains limited and are invisible to the clinician except in the rare instances where a kidney biopsy is obtained. Traditional markers such as estimated glomerular filtration rate (eGFR) and albuminuria reflect downstream consequences of kidney damage but do not directly capture microvascular health.

Recent advances in optical technologies including laser Doppler flowmetry (LDF) have enabled noninvasive assessment of microvascular function in peripheral tissues.^6,7^ LDF and its response to local heating quantifies skin microvascular blood flow, reflecting arteriolar and venular perfusion.^8^ Local skin heating induces arteriolar vasodilation, and the magnitude of the perfusion increase reflects endothelial-dependent reactive hyperemia.^9^ However, the extent to which peripheral microvascular measures captured by LDF relate to established markers of kidney function and structural kidney damage remains poorly defined.^10^

In the present study, we hypothesized that skin microvascular blood flow responses to heat-induced hyperemia are impaired in CKD and that LDF-derived measures may serve as accessible, noninvasive surrogate markers of kidney function and systemic microvascular health. To test this hypothesis, we examined the relationships between both resting and heat-induced changes in skin perfusion and markers of kidney function among individuals with established CKD. In a subset of participants who underwent clinically indicated kidney biopsies, we additionally examined associations between LDF-derived measures and histologic markers of kidney damage, including glomerulosclerosis (GS) and interstitial fibrosis/tubular atrophy (IFTA). By integrating noninvasive microvascular phenotyping with functional and histologic markers, this study aims to define the potential role of LDF as a translational tool for assessing systemic microvascular health in CKD.

## Methods

### Study design and population

The Microvascular Assessment in Patients with CKD (MAP-CKD) study is a cross-sectional investigation of individuals with CKD recruited from nephrology clinics at the University of California, San Diego (UCSD) between January 2023 and December 2025. Eligible participants had an estimated glomerular filtration rate (eGFR) <90 mL/min/1.73 m² as defined by the CKD-EPI equation.^11^ Proteinuria was assessed using a spot urine protein-to-creatinine ratio (uPCR) and categorized as normal to mildly increased (<0.15 mg/mg), moderately increased (0.15-0.5 mg/mg), or severely increased (>0.5 mg/mg).^12^ Exclusion criteria included age <18 years, pregnancy, ESKD requiring dialysis, prior kidney transplantation, and inability or unwillingness to provide consent. A subset of recruited participants underwent kidney biopsies for various clinical indications among whom we obtained kidney biopsy reports and assessed the degree of GS and IFTA. The study was approved by the UCSD Institutional Review Board, and all participants provided written informed consent.

### Laser Doppler flowmetry measurement

LDF was used to quantify cutaneous microvascular perfusion of the forearm during a standardized local thermal hyperemia protocol (PF 457, Perimed, Stockholm, Sweden).^13^ Measurements were performed in a quiet, temperature-controlled room, 24°C, following a 20-minute acclimatization period, with participants resting in a supine position to minimize sympathetic activation and temperature-related fluctuations in blood flow. The probe was placed on the ventral surface of the non-dominant forearm, approximately 10 cm proximal to the wrist while avoiding visible veins, and stable signal quality was verified before acquisition.

The protocol consisted of a 20-minute baseline phase at ∼30°C followed by local heating to 44°C for an additional 20 minutes to induce maximal vasodilation. Continuous perfusion signals were recorded in perfusion units (PU) representing the product of red blood cell velocity and concentration and exported as time-series data for quantitative analysis. For each recording (baseline and maximal), the most stable 5–10-minute baseline and hyperemic segments were automatically identified using a rolling-window algorithm that minimized temporal variance of smoothed PU values. Baseline PU was defined as the median PU during the stable period at ∼30 °C, and maximal PU as the median PU during the stable period at 44°C for eat least 5 minutes. The percentage change in perfusion units (%ΔPU) was calculated as: %ΔPU = ((maximal PU − baseline PU) / baseline PU) × 100, and used to define microvascular functional reserve, reflecting the capacity of the microvasculature to augment blood flow in response to thermal stimulation.

### Histopathology assessment

A standardized semi-quantitative assessment of GS and IFTA was performed on trichrome-stained biopsy specimens from a subset of CKD participants (n=20) who underwent kidney biopsy for clinical indications. LDF measurements were obtained within 30 days of kidney biopsy in all participants. A board-certified kidney nephropathologist who was blinded to clinical and imaging data assigned histologic scores for GS and IFTA using validated criteria.^14^ GS was defined as global or segmental collapse of glomerular capillary walls and consolidation of the glomerular tuft by extracellular matrix, resulting in capillary luminal obliteration. IFTA was assessed by the degree of renal cortex involved by established fibrosis; and tubular atrophy expressed as a percentage. To ensure accurate representation and reproducibility, at least 10 glomeruli were scored for each participant.

### Statistical analysis

Categorical variables were summarized as frequencies and percentages, and continuous variables were presented as means with standard deviations (SD) or medians with interquartile ranges (IQR), as appropriate. Normality was assessed using the Shapiro–Wilk test. Spearman’s rank correlation coefficients were used to evaluate the relationship between log-transformed LDF measures and kidney function and histopathology including eGFR, log-transformed uPCR, and percentage GS and IFTA. Differences in skin perfusion across CKD stages were assessed using the Kruskal-Wallis test with Dunn’s post hoc correction for multiple comparisons.

Principal component analysis (PCA) was performed on z-score standardized LDF measures (baseline PU and microvascular functional reserve) to assess the underlying dimensional structure of microvascular perfusion. Baseline PU was reverse-coded so that higher values consistently reflected more favorable microvascular function. The first principal component explained 84% of the total variance and represented a shared axis of microvascular functional reserve, integrating both resting perfusion and heat-induced response. Unsupervised k-means clustering (k=4) was then applied to standardized LDF variables to identify data-driven microvascular phenotypes.

Multivariable linear regression was used to evaluate associations of microvascular perfusion with eGFR and log-transformed uPCR, adjusting for age, sex, race, body mass index (BMI), diabetes, hypertension, peripheral vascular disease, and cardiovascular disease (CVD). A two-tailed P-value <0.05 was considered statistically significant. All analyses were conducted using R version 4.2.2, and figures were generated in GraphPad Prism version 10.0.0.

## Results

The study included 150 participants with CKD, with a mean (SD) age of 64 (14); 46% were female, and 45% White, 22% Hispanic, 19% Asian, and 13% African American (**Table 1**). The mean eGFR was 42 (21) ml/min/1.73 m^2^, with 20% of participants in stage 2, 49% in stage 3, 21% in stage 4, and 10% in stage 5. The median (IQR) urine protein-to-creatinine ratio (uPCR) was 0.21 (0.11 to 1.20) mg/mg. The most common comorbidities were hypertension (83%), diabetes (38%), and CVD (40%).

**Table 1.** Characteristics of the participants in the MAP-CKD study.

| Characteristics | Overall |
| --- | --- |
| Number | 150 |
| Demographics |  |
| Age, mean (*SD*) | 64 (14) |
| Female, n (%) | 69 (46) |
| Race, n (%) |  |
| Asian/Pacific Islander | 28 (19) |
| African American | 19 (13) |
| Hispanic | 33 (22) |
| White | 68 (45) |
| BMI, mean (*SD*) (kg/m<sup>2</sup>) | 27 (6) |
| Medical history, n (%) |  |
| Diabetes | 57 (38) |
| Hypertension | 125 (83) |
| Heart disease | 60 (40) |
| Peripheral vascular disease | 34 (22) |
| Autoimmune | 26 (17) |
| Medication use, n (%) |  |
| ACE/ARB inhibitor | 99 (66) |
| GLP-1 agonist | 15 (10) |
| SGLT-2 inhibitor | 54 (36) |
| Chemotherapy | 20 (13) |
| Laboratory data |  |
| Serum creatinine (mg/dl), median (IQR) | 1.6 (1.3 to 2.5) |
| eGFR (mL/min/1.73 m<sup>2</sup>), mean (*SD*) | 42 (21) |
| Urine PCR (mg/mg), median (IQR) | 0.21 (0.11 to 1.2) |
| CKD stage, n (%) |  |
| Stage 2 | 30 (20) |
| Stage 3a | 32 (22) |
| Stage 3b | 41 (27) |
| Stage 4 | 32 (21) |
| Stage 5 | 15 (10) |

Abbreviations: BMI, body mass index; ACE/ARB, angiotensin-converting enzyme inhibitors/angiotensin receptor blockers; GLP-1, glucagon-like peptide-1; SGLT-2, sodium/glucose cotransporter 2; Urine PCR, urine protein-to-creatinine ratio; eGFR, estimated glomerular filtration rate; SD, standard deviation; IQR, interquartile range; CKD, chronic kidney disease.

The median (IQR) baseline PU was 10 (6 to 16), maximal PU was 75 (46 to 118), and percent PU change was 677% (262 to 1504). Higher baseline PU was associated with lower eGFR (r = -0.17, p = 0.03) and increased progressively with advanced CKD stage (p for trend =0.03) (**Figure 1A-B**, **Table 2**). In unadjusted linear regression models, baseline PU was inversely associated with eGFR (β = -12; 95% CI, -24 to -1), with a stronger association after adjustment for demographics and clinical covariates including proteinuria (β = -18; 95% CI, -29 to -6) (**Table 3**). Greater percent change in PU was associated with higher eGFR (r = 0.21, p < 0.01) and decreased with advancing CKD stage (p for trend <0.01) (**Figure 1C-D**, **Table 2**). In regression analyses, greater percent change in PU was associated with higher eGFR in the unadjusted model and remained significantly associated after multivariable adjustment (β = 7; 95% CI, 2 to 12) (**Table 3**). Sensitivity analysis further adjusting for baseline PU did not attenuate this association (β = 7; 95% CI, 1 to 14) (**Supplemental Table 1**).

**Figure 1.**
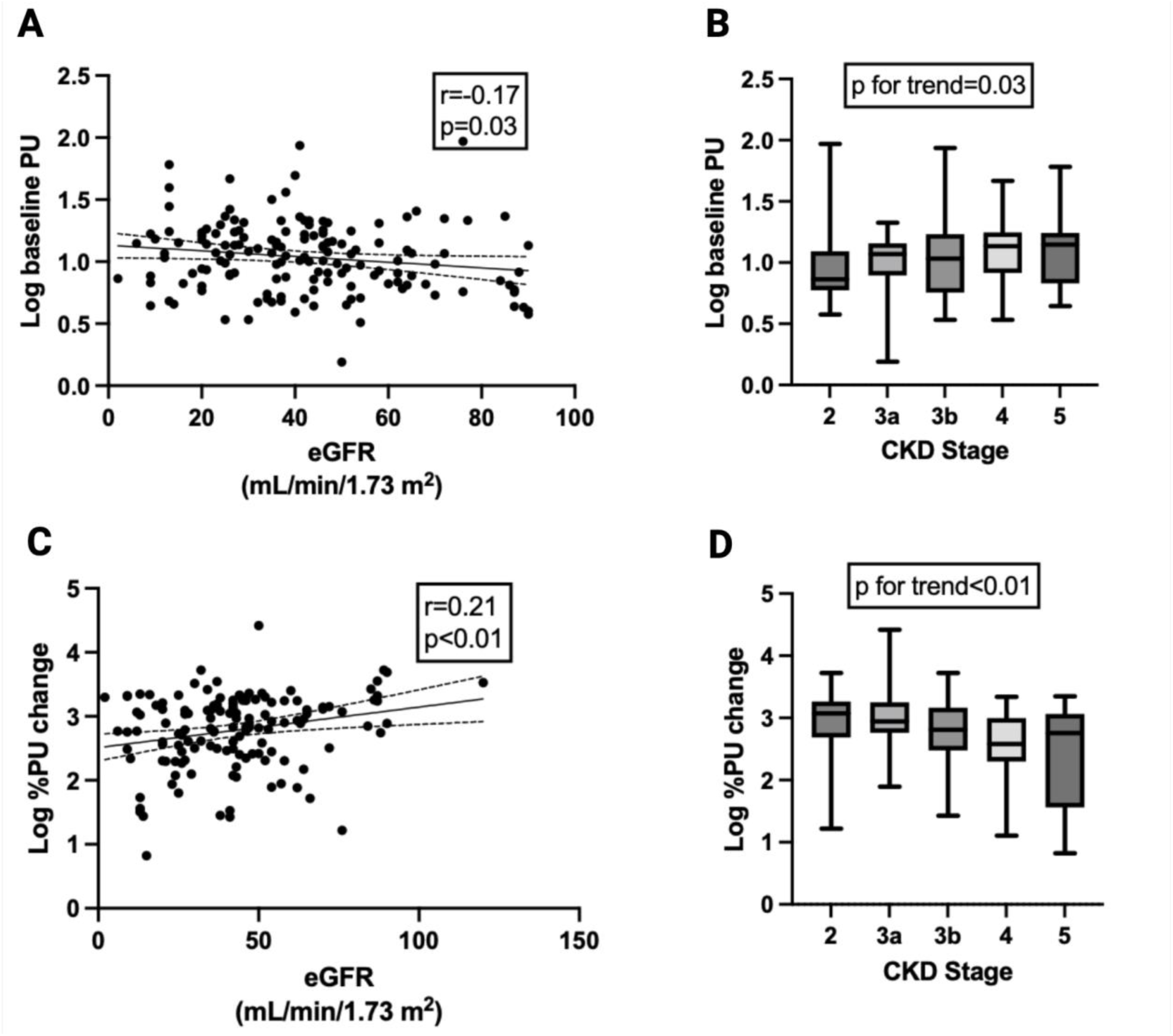
The association of LDF-derived measures with eGFR and CKD stage severity in CKD. The correlation between baseline PU and A) eGFR and B) CKD stage, and between %PU change and C) eGFR and D) CKD stage. Spearman correlation coefficients were used to estimate the univariate relationship between LDF measures and eGFR. The box plots represent median and IQR, and the whiskers represent minimum and maximum values.

**Table 2.** LDF-derived measures across CKD stages.

| **Microvascular**<br>**function measures** | **Overall**<br>**(n=150)** | **CKD 2**<br>**(n=30)** | **CKD 3a**<br>**(n=32)** | **CKD 3b**<br>**(n=41)** | **CKD 4**<br>**(n=32)** | **CKD 5**<br>**(n=15)** |
| --- | --- | --- | --- | --- | --- | --- |
| Baseline PU,<br>median (IQR) | 10 (6 to 16) | 7 (5 to 11) | 11 (7 to 13) | 10 (5 to 17) | 13 (8 to 17) | 13 (6 to 16) |
| Maximal PU,<br>median (IQR) | 75 (46 to 118) | 89 (49 to 131) | 93 (62 to 167) | 75 (47 to 117) | 53 (41 to 91) | 67 (45 to 92) |
| % PU change,<br>median (IQR) | 677 (262 to 1504) | 1182 (496 to 1849) | 884 (575 to 1780) | 649 (301 to 1464) | 381 (198 to 1008) | 569 (36 to 1160) |

Abbreviations: IQR, interquartile ranges; PU, perfusion unit; CKD, chronic kidney disease; LDF, laser doppler flowmetry.

**Table 3.** The association of LDF measures with eGFR and urine PCR in CKD.

| **Covariate adjustment** | **Baseline PU vs eGFR** |  | **%PU change vs eGFR** |  |
| --- | --- | --- | --- | --- |
|  | Beta (95% CI) | P-value | Beta (95% CI) | P-value |
| Model 1 (unadjusted) | -12 (-24 to -1) | 0.04 | 7 (2 to 13) | <0.01 |
| Model 2 | -15 (-27 to -3) | 0.01 | 8 (2 to 13) | <0.01 |
| Fully adjusted model | -18 (-29 to -6) | <0.01 | 7 (2 to 12) | <0.01 |
|  | **Baseline PU vs uPCR** |  | **%PU change vs uPCR** |  |
|  | Beta (95% CI) | P-value | Beta (95% CI) | P-value |
| Model 1 (unadjusted) | -0.09 (-0.25 to 0.07) | 0.28 | -0.05 (-0.13 to 0.03) | 0.20 |
| Model 2 | -0.09 (-0.23 to 0.05) | 0.19 | -0.01 (-0.08 to 0.05) | 0.65 |
| Fully adjusted model | -0.15 (-0.28 to -0.01) | 0.03 | 0.01 (-0.05 to 0.07) | 0.73 |

$\beta$ coefficients represent the change in eGFR or Log uPCR per one-log unit increase in each LDF measure. per one-log unit increase in each LDF measure. Model 1 is unadjusted. Model 2 includes adjustments for age, sex, race, diabetes, hypertension, BMI, peripheral vascular disease, and CVD. The fully adjusted model further adjusts for uPCR or eGFR. Abbreviations: LDF, laser doppler flowmetry; PU, perfusion unit; eGFR, estimated glomerular filtration rate; BMI, body mass index; uPCR, urine protein creatinine ratio.

Baseline PU was not associated with uPCR, and no meaningful differences in baseline PU was observed across proteinuria categories (**Figure 2A-B**). Consistently, baseline PU showed no association with uPCR in unadjusted or partially adjusted models. After additional adjustment for eGFR, baseline PU was inversely associated with uPCR (β = -0.15; 95% CI, -0.28 to -0.01) (**Table 3**). Percent change in PU was also not correlated with uPCR and did not differ across proteinuria categories (**Figure 2C-D**). In unadjusted models, lower percent change in PU was marginally associated with higher uPCR, but this association was attenuated after partial and full adjustment (**Table 3**).

**Figure 2.**
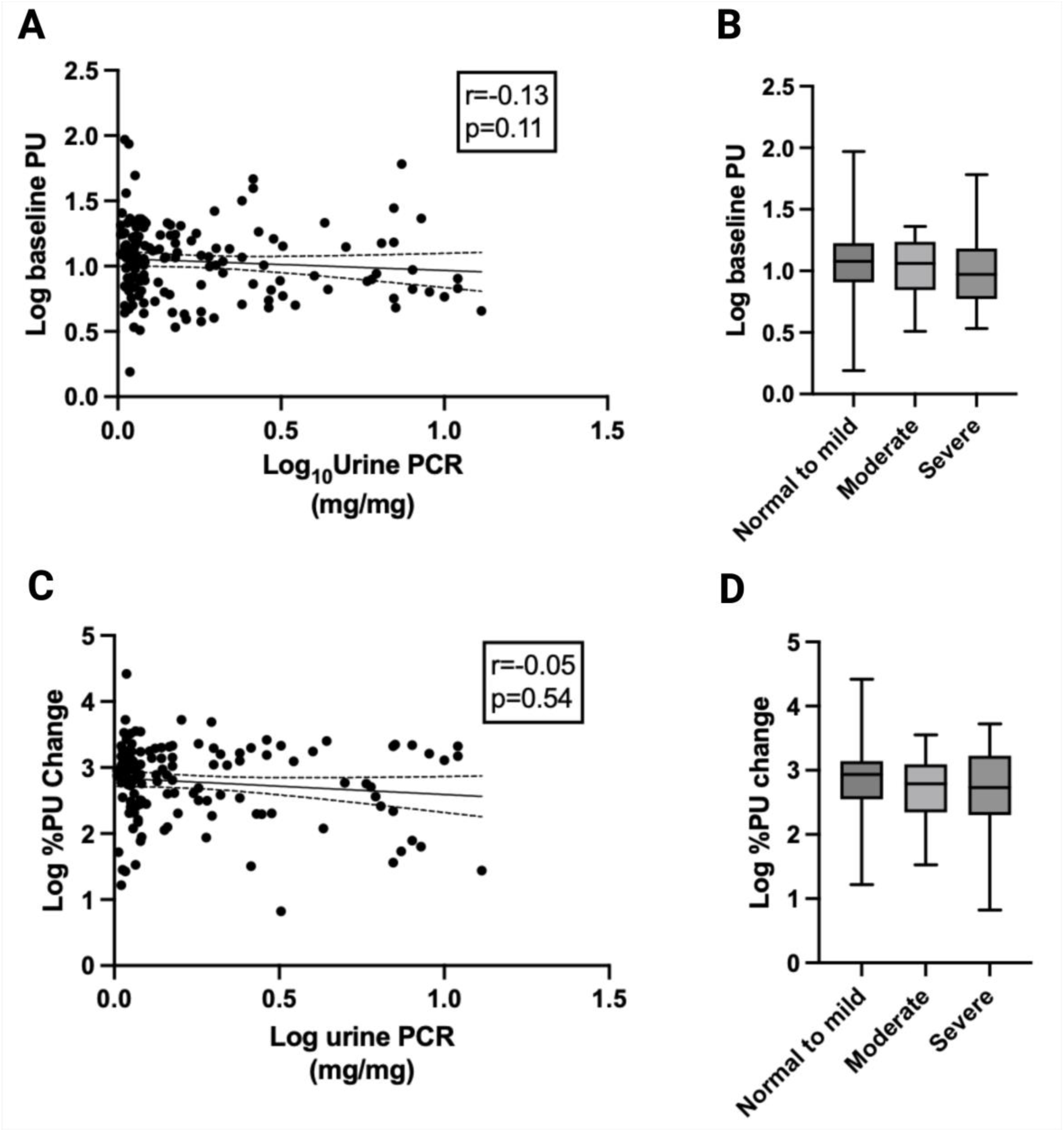
The Association of LDF-derived measures and proteinuria severity in CKD. The relationship between baseline PU and A) uPCR and B) proteinuria severity; and between %PU change and C) uPCR and D) proteinuria severity. Spearman correlation coefficients were used to estimate the univariate relationship between LDF measures and proteinuria. The box plots represent median and IQR, and the whiskers represent minimum and maximum values.

Unsupervised clustering of LDF perfusion measures identified four distinct microvascular phenotypes defined by differences in resting PU and microvascular functional reserve (**Figure 3**). Patient-level demographic and clinical characteristics across LDF-derived phenotypes are summarized in **Supplemental Table 2**. Cluster 3 (n = 13) represented the most impaired phenotype, characterized by the highest resting PU, lowest percent change in PU, and the lowest mean eGFR (**Figure 3 and Supplemental Table 3)**. Cluster 4 (n = 39) exhibited moderate microvascular impairment, with higher resting PU, reduced percent change in PU, and lower mean eGFR. In contrast, Clusters 1 (n = 54) and 2 (n = 44) demonstrated more preserved microvascular function with lower resting PU and greater percent change in PU. Cluster 2 exhibited the most favorable profile, with the lowest resting perfusion, highest percent change in PU, and highest mean eGFR, whereas Cluster 1 showed intermediate values across these measures (**Figure 3 and Supplemental Table 3)**.

**Figure 3.**
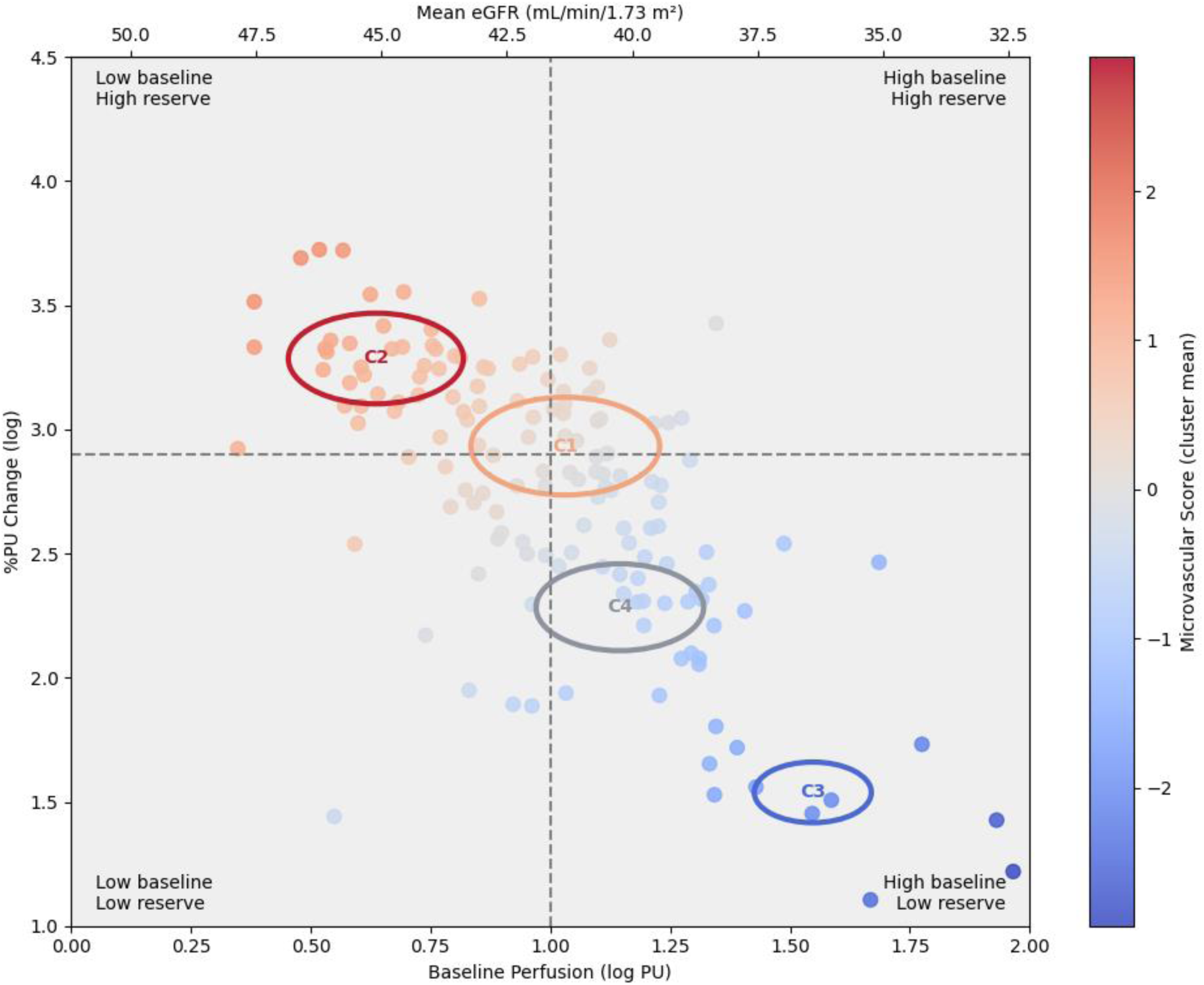
LDF-derived microvascular phenotypes in CKD identified using unsupervised clustering (k=4). Baseline perfusion and heat-induced perfusion response were used for k-means clustering. Circles represent cluster centroids scaled by sample size, and colors indicate cluster-level mean microvascular health scores.

Baseline characteristics of the participants with available kidney biopsy are shown in **Supplemental Table 4**. Compared with the overall CKD cohort, these participants were younger, had a higher prevalence of autoimmune diseases, higher levels of uPCR, and higher eGFR. The most common biopsy diagnoses were lupus nephritis (35%), diabetic nephropathy (15%), and focal segmental glomerulosclerosis (FSGS) (10%) (**Supplemental Table 4**). Among the biopsy participants, higher baseline PU was associated with greater GS (r=0.59) and IFTA (r=0.57) percentages (**Figure 4A-B**). Conversely, greater percent change in PU was associated with lower GS (r=-0.50) and IFTA (r=-0.58) (**Figure 4C-D**).

**Figure 4.**
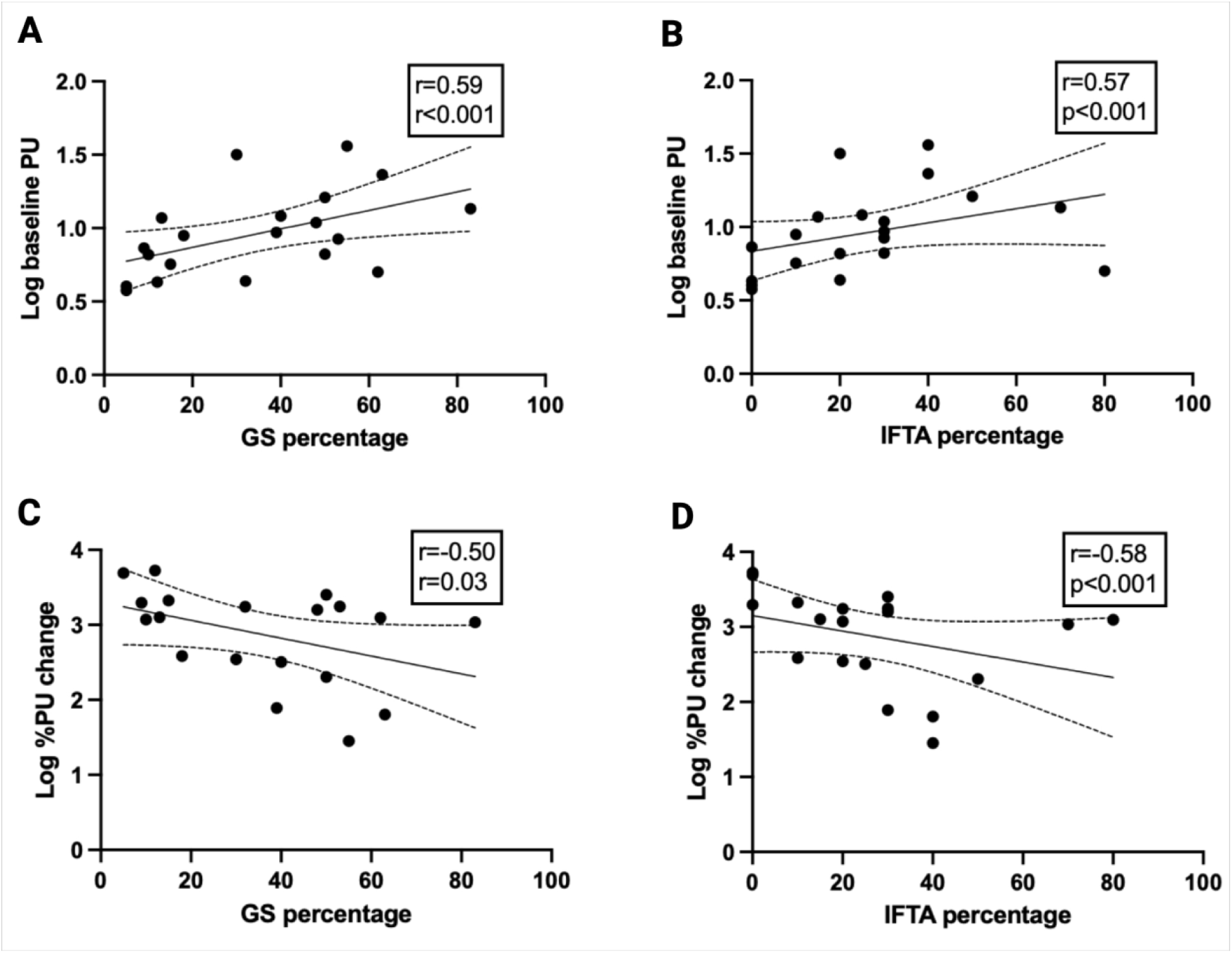
The relationship between LDF-derived measures and histopathology results in the subset of MAP-CKD participants with kidney biopsy (n=20). The relationship between baseline PU with A) GS percentage, B) IFTA percentage, and the relationship between percent PU change with C) GS percentage and D) IFTA percentage. Spearman correlation coefficients were used to estimate the univariate relationship between LDF measures and histopathology severity.

## Discussion

In this study, we examined the associations of microvascular blood flow in the skin, assessed by LDF, with kidney function (eGFR), uPCR, and histopathology in patients with CKD. Lower eGFR was associated with higher baseline PU and a reduced percent change in PU in response to heat induced hyperemia. These associations remained significant even after adjustment for demographics and clinical covariates. In contrast, uPCR was not associated with either baseline PU or percent change in PU. Unsupervised clustering of perfusion measures identified four distinct microvascular phenotypes with graded differences in resting perfusion and functional reserve (defined by percent change in PU). The associations of baseline PU and % change in PU with kidney function biomarkers were largely confirmed by histopathology among the subset with kidney biopsies. Among these individuals, higher baseline PU and lower percent change in PU were associated with greater severity of GS and IFTA, respectively. Taken together, these findings support a framework in which microvascular dysfunction in CKD is characterized not solely by alterations in resting perfusion, but also by impairment in microvascular functional reserve. In this context, more advanced CKD was associated with a pattern of elevated resting perfusion accompanied by reduced capacity to augment flow, reflecting impaired microvascular adaptability to physiologic stress.

LDF has been used to assess microvascular function in conditions such as hypertension, insulin resistance, diabetic nephropathy, and systemic sclerosis in prior studies.^7,15–17^ In the present study, we extend its application to CKD by demonstrating associations between peripheral microvascular perfusion with kidney function and histopathologic injury. At first glance, the observed association between more advanced CKD and higher resting perfusion may appear counterintuitive, as microvascular dysfunction is often conceptualized as reduced tissue blood flow. However, elevated baseline perfusion in this context may reflect impaired autoregulatory control and compensatory vasodilation rather than preserved microvascular integrity. The limited percent recruitment in response to the heat stimulus is consistent with this hypothesis as well. This is in line with previous studies showing wider retinal arterioles and increased resting blood flux in in persons with diabetes.^18,19^ Chronic inflammation, oxidative stress, and altered local metabolic signaling may promote sustained basal vasodilation leading to elevated resting perfusion while simultaneously constraining the capacity for further vasodilatory recruitment.^20^ These findings were associated with glomerulosclerosis on kidney biopsy. It may be that capillary damage and drop-out in the most distal aspects of the circulatory tree (aka “rarefaction”) may promote redistribution of blood flow through a reduced number of dilated peripheral vessels, resulting in higher measured perfusion in the skin.

Importantly, the observed reduction in percentage change in perfusion cannot be explained solely by elevated baseline perfusion. Even after accounting for baseline differences, the relationship between CKD severity and impaired vasodilatory response persisted, suggesting an intrinsic defect in microvascular reactivity. The uremic milieu, endothelial dysfunction, reduced nitric oxide bioavailability, and vascular remodeling may have impaired the ability of the microvasculature to appropriately augment blood flow in response to physiologic stimuli.^21^ Overall, these findings suggest that peripheral microvascular abnormalities reflect broader systemic vascular alterations relevant to the pathophysiology of CKD.

In contrast to some prior studies, we did not observe an association between LDF-derived measures and uPCR.^22,23^ This difference likely reflects variation in study populations and underlying disease burden and mechanisms. Whereas proteinuria primarily reflects microvascular damage to the glomerular filtration barrier, LDF-derived measures capture dynamic endothelial and microvascular vasomotor function.^24^ In advanced CKD, these processes may become partially dissociated, with structural glomerular damage driving protein excretion while systemic endothelial dysfunction and microvascular rarefaction impair vascular reactivity.^25–27^ In addition, therapies such as renin–angiotensin system inhibitors and sodium-glucose transporter (SGLT2) inhibitors may reduce proteinuria without fully restoring endothelial or microvascular function. These findings suggest that LDF-derived measures reflect a complementary dimension of microvascular health that is not captured by proteinuria alone.

Unsupervised clustering further revealed distinct microvascular phenotypes, highlighting heterogeneity in vascular tone and microvascular functional reserve among individuals with CKD. These findings suggest that microvascular dysfunction in CKD comprises distinct functional states that are not fully captured by conventional markers such as eGFR. This phenotype-based framework underscores the importance of assessing dynamic vascular responses in addition to resting perfusion and provides a basis for improved risk stratification and identification of patient subgroups with differing degrees of vascular adaptability. These findings also raise the hypothesis that impairment of microvascular functional reserve may precede or accompany decline in kidney function and progression of fibrotic injury. Distinct microvascular phenotypes may therefore identify patients with differing risk profiles or therapeutic responsiveness, a hypothesis that warrants evaluation in prospective studies. Overall, this functional phenotyping approach provides a framework for understanding microvascular dysfunction in CKD and may inform future strategies aimed at improving vascular function and modifying disease progression.

Although limited to a subset of 20 participants with kidney biopsy data, the inclusion kidney biopsies represents a key strength of this study. This enabled the examination of associations between LDF-derived microvascular measures and glomerular and interstitial injury. And it provides preliminary evidence of the potential value of non-invasive LDF assessment for underlying structural kidney damage in patients with CKD. Prior imaging studies in CKD have demonstrated that measures such as cortical blood volume assessed by CT angiography and retinal choroidal thickness measured by optical coherence tomography may reflect underlying kidney damage; however, these approaches are either invasive or not widely accessible.^28^ Thus, our findings suggest that LDF measures are not only associated with CKD severity but may also reflect underlying kidney histopathologic injury. Whether these associations are independent of demographic factors and comorbid conditions requires further confirmation in larger cohorts.

Additional strengths of this study include the use of non-invasive LDF with a validated protocol, enabling assessment of both resting and dynamic aspects of microvascular function. We applied an unsupervised clustering approach to characterize heterogeneity in LDF-derived microvascular function across individuals with CKD. In addition, the study integrated clinical, laboratory, and histopathologic data, allowing for a comprehensive evaluation of how baseline skin blood flow and dynamic changes in response to heat relate to kidney function and structural injury. The ability to link peripheral microvascular measures with biopsy features such as GS and IFTA adds biological insight to the observed associations. Finally, the study cohort was diverse, with representation across racial and ethnic groups and CKD stages, enhancing the generalizability of our findings to broader clinical populations.

The study also has important limitations. First, the cross-sectional design precludes causal inference and assessment of temporal relationships between impaired microvascular perfusion and CKD progression. We plan longitudinal follow-up of this cohort to evaluate whether microvascular measures predict future decline in kidney function and adverse clinical outcomes, thereby addressing this limitation. Second, although the overall sample size was modest, the cohort included individuals across all CKD stages which may support generalizability across spectrum of disease severity. In addition, while some studies have decomposed skin blood flow into multiple contributing domains, including endothelial, neurogenic, myogenic, respiratory, and cardiac components, our measurements reflect an integrated signal of skin perfusion.^28^ Although heat-induced blood flow is largely endothelium-dependent, the present analysis does not distinguish endothelium-dependent from endothelium-independent vasodilatory mechanisms.^29^

In conclusion, CKD is associated with elevated resting perfusion and impaired microvascular functional reserve in the skin, consistent with systemic microvascular dysfunction. LDF-derived microvascular phenotypes reveal physiologic heterogeneity that is not captured by traditional clinical markers. Moreover, they may help inform risk stratification and the targeting of therapies aimed at improving microvascular function.

## Data Availability

The data that support the findings of this study are available from the corresponding author upon request. De-identified individual participant data, the study protocol, and statistical analysis code may be shared for academic purposes, subject to institutional data use agreements and IRB approval where applicable.

## Author Contributions

The study was conceptualized by Rakesh Malhotra (RM), Alfons J.H.M. Houben (AHH), and Joachim H. Ix (JHI). Methodology development was performed by RM, AHH, and JHI. Data acquisition was conducted by Jason Yang (JY) and Basma Ghanim (BG) and data curation was carried out by Armin Ahmadi (AA), Masfiqur Rahaman (MR), Amol Harsh (AH), Subhasis Dasgupta (SD), and RM. Formal analysis was conducted by AA. The original draft of the manuscript was written by RM and AA. Tauhidur Rahman (TR) and Robert N. Weinreb (RNW) provided guidance on imaging interpretation and critical revision of the manuscript. Supervision was provided by RM and JHI. Funding acquisition was led by RM and JHI. All authors reviewed and edited the manuscript and approved the final version for submission.

---

**Supplemental Table 1.** Sensitivity analysis adjusting for baseline perfusion showing the association of percent PU change with eGFR and proteinuria.

| Model | %PU change vs eGFR |  |
| --- | --- | --- |
|  | Beta (95% CI) | P-value |
| Fully adjusted model | 7 (1 to 14) | 0.04 |
|  | %PU change vs uPCR |  |
|  | Beta (95% CI) | P-value |
| Fully adjusted model | -0.00 (-0.04 to 0.03) | 0.92 |

$\beta$ coefficients represent the change in eGFR or Log uPCR per one-log unit increase in percent PU change. The fully adjusted model adjustments for age, sex, race, diabetes, hypertension, BMI, peripheral vascular disease, CVD, uPCR/eGFR, and baseline PU. Abbreviations: LDF, laser doppler flowmetry; PU, perfusion unit; eGFR, estimated glomerular filtration rate; BMI, body mass index; uPCR, urine protein creatinine ratio.

**Supplemental table 2.** Patient characteristics stratified by LDF-derived microvascular health score.

| Characteristic | Cluster 1<br>(n=54) | Cluster 2<br>(n=44) | Cluster 3<br>(n=13) | Cluster 4<br>(n=39) |
| --- | --- | --- | --- | --- |
| Age, mean (SD) | 67 (12) | 59 (16) | 60 (17) | 68 (12) |
| Female (%) | 21 (39) | 24 (55) | 5 (38) | 18 (46) |
| BMI, mean (SD) | 27 (6) | 28 (7) | 26 (4) | 25 (4) |
| Diabetes (%) | 17 (31) | 17 (39) | 5 (38) | 18 (46) |
| Hypertension (%) | 47 (87) | 32 (73) | 11 (85) | 35 (90) |
| Cardiovascular disease (%) | 22 (41) | 15 (34) | 1 (8) | 22 (56) |
| Peripheral vascular disease (%) | 10 (19) | 10 (23) | 7 (54) | 7 (18) |
| Autoimmune disease (%) | 8 (15) | 10 (23) | 2 (15) | 5 (13) |
| ACE/ARB (%) | 36 (67) | 34 (77) | 5 (38) | 25 (64) |
| SGLT2 inhibitor (%) | 21 (39) | 16 (36) | 4 (31) | 14 (36) |
| GLP-1 agonist (%) | 8 (15) | 4 (9) | 0 (0) | 3 (8) |
| Serum creatinine, median (IQR) | 1.60 (1.21-2.27) | 1.44 (1.10-1.82) | 1.70 (1.30-3.40) | 1.80 (1.40-2.65) |
| eGFR, mean (SD) | 43 (20) | 47 (24) | 37 (23) | 37 (16) |
| uPCR, median (IQR) | 0.14 (0.09-0.50) | 0.45 (0.11-2.01) | 0.16 (0.07-2.20) | 0.25 (0.17-0.95) |
| CKD stage, n (%) |  |  |  |  |
| Stage 2 | 10 (18) | **13 (30)** | 3 (23) | 4 (10) |
| Stage 3a | **15 (28)** | 10 (23) | 0 (0) | 7 (18) |
| Stage 3b | 13 (24) | 13 (30) | **4 (31)** | 11 (28) |
| Stage 4 | 11 (20) | 5 (11) | 2 (15) | **14 (36)** |
| Stage 5 | 5 (10) | 3 (6) | **4 (31)** | 3 (8) |
| LDF-derived Score | 0.1 | 1.0 | -1.9 | -0.6 |

Abbreviations: BMI, body mass index; ACE/ARB, angiotensin-converting enzyme inhibitors/angiotensin receptor blockers; GLP-1, Glucagon-like peptide-1; SGLT-2, sodium/glucose cotransporter 2; Urine PCR, Urine protein-to-creatinine ratio; eGFR, estimated glomerular filtration rate; SD, standard deviation; IQR, interquartile range; CKD, chronic kidney disease.

**Supplemental Table 3.** The distribution of LDF-derived measurements and eGFR across LDF-based scores in CKD.

| Cluster | n | %PU change | Baseline PU | eGFR | LDF Score |
| --- | --- | --- | --- | --- | --- |
| 1 | 54 | 2.9 | 1.1 | 43 | 0.1 |
| 2 | 44 | 3.3 | 0.7 | 47 | 1.0 |
| 3 | 13 | 1.5 | 1.5 | 37 | -1.9 |
| 4 | 39 | 2.3 | 1.2 | 37 | -0.6 |

**Supplemental Table 4.** Characteristics of the MAP CKD participants with available kidney biopsy (n=20)

| Characteristics | Overall | CKD with biopsy |
| --- | --- | --- |
| Number | 150 | 20 |
| Demographics |  |  |
| Age, mean (*SD*) | 64 (14) | 53 (17) |
| Female, n (%) | 69 (46) | 12 (60) |
| Race, n (%) |  |  |
| Asian/Pacific Islander | 29 (19) | 8 (40) |
| African American | 19 (13) | 2 (10) |
| Hispanic | 33 (22) | 3 (15) |
| White | 68 (45) | 7 (35) |
| BMI, mean (*SD*) (kg/m<sup>2</sup>) | 27 (6) | 26 (5) |
| Biopsy findings, n (%) |  |  |
| Lupus |  | 7 (35) |
| Diabetic nephropathy |  | 3 (15) |
| FSGS |  | 2 (10) |
| Other |  | 7 (35) |
| Medical history and lifestyle, n (%) |  |  |
| Diabetes | 57 (38) | 7 (35) |
| Hypertension | 125 (83) | 14 (70) |
| Cardiovascular disease | 60 (40) | 5 (25) |
| Peripheral vascular disease | 34 (22) | 2 (10) |
| Autoimmune | 26 (17) | 12 (60) |
| Medication use, n (%) |  |  |
| ACE/ARB inhibitor | 99 (66) | 15 (75) |
| GLP-1 agonist | 15 (10) | 1 (5) |
| SGLT-2 inhibitor | 54 (36) | 10 (50) |
| Chemotherapy | 20 (13) | 3 (15) |
| Laboratory data |  |  |
| Serum creatinine (mg/dl), median (IQR) | 1.6 (1.3 to 2.5) | 1.4 (0.9 to 1.6) |
| Urine PCR (mg/mg), median (IQR) | 0.21 (0.1 to 1.2) | 1.4 (0.8 to 2.8) |
| eGFR (mL/min/1.73 m<sup>2</sup>), mean (*SD*) | 42 (21) | 53 (27) |
| Baseline PU, median (IQR) | 10 (6 to 16) | 8 (4 to 12) |
| Percent PU change, median (IQR) | 677 (262 to 1504) | 1243 (319 to 1979) |

Abbreviations: BMI, body mass index; FGSS, Focal segmental glomerulosclerosis; ACE/ARB angiotensin-converting enzyme inhibitors/angiotensin receptor blockers; GLP-1, Glucagon-like peptide-1; SGLT-2, sodium/glucose cotransporter 2; Urine PCR, Urine protein-to-creatinine ratio; eGFR, estimated glomerular filtration rate; SD, standard deviation; IQR, interquartile range.

## References

1. Bidani AK, Polichnowski AJ, Loutzenhiser R, et al. Renal microvascular dysfunction, hypertension and CKD progression. Curr Opin Nephrol Hypertens. Jan 2013;22(1):1–9. doi:10.1097/MNH.0b013e32835b36c1

2. Afsar B, Afsar RE, Dagel T, et al. Capillary rarefaction from the kidney point of view. Clin Kidney J. Jun 2018;11(3):295–301. doi:10.1093/ckj/sfx133

3. Stam F, van Guldener C, Becker A, et al. Endothelial dysfunction contributes to renal function-associated cardiovascular mortality in a population with mild renal insufficiency: the Hoorn study. J Am Soc Nephrol. Feb 2006;17(2):537–45. doi:10.1681/asn.2005080834

4. Lilien MR, Groothoff JW. Cardiovascular disease in children with CKD or ESRD. Nat Rev Nephrol. Apr 2009;5(4):229–35. doi:10.1038/nrneph.2009.10

5. Domsic RT, Dezfulian C, Shoushtari A, et al. Endothelial dysfunction is present only in the microvasculature and microcirculation of early diffuse systemic sclerosis patients. Clin Exp Rheumatol. Nov-Dec 2014;32(6 Suppl 86):S-154–60.

6. Antonios TF, Rattray FE, Singer DR, et al. Maximization of skin capillaries during intravital video-microscopy in essential hypertension: comparison between venous congestion, reactive hyperaemia and core heat load tests. Clin Sci (Lond). Oct 1999;97(4):523–8.

7. Allen J, Howell K. Microvascular imaging: techniques and opportunities for clinical physiological measurements. Physiol Meas. Jul 2014;35(7):R91–r141. doi:10.1088/0967-3334/35/7/r91

8. Low DA, Jones H, Cable NT, et al. Historical reviews of the assessment of human cardiovascular function: interrogation and understanding of the control of skin blood flow. Eur J Appl Physiol. Jan 2020;120(1):1–16. doi:10.1007/s00421-019-04246-y

9. Cheng JL, MacDonald MJ. Effect of heat stress on vascular outcomes in humans. J Appl Physiol (1985). Mar 1 2019;126(3):771–781. doi:10.1152/japplphysiol.00682.2018

10. Babos L, Járai Z, Nemcsik J. Evaluation of microvascular reactivity with laser Doppler flowmetry in chronic kidney disease. World J Nephrol. Aug 6 2013;2(3):77–83. doi:10.5527/wjn.v2.i3.77

11. Inker LA, Eneanya ND, Coresh J, et al. New Creatinine- and Cystatin C–Based Equations to Estimate GFR without Race. New England Journal of Medicine. 2021;385(19):1737–1749. doi:doi:10.1056/NEJMoa2102953

12. KDIGO 2024 Clinical Practice Guideline for the Evaluation and Management of Chronic Kidney Disease. Kidney Int. Apr 2024;105(4s):S117–s314. doi:10.1016/j.kint.2023.10.018

13. Freccero C, Holmlund F, Bornmyr S, et al. Laser doppler perfusion monitoring of skin blood flow at different depths in finger and arm upon local heating. Microvascular Research. 2003/11/01/ 2003;66(3):183–189. 10.1016/j.mvr.2003.06.001

14. Sethi S, D’Agati VD, Nast CC, et al. A proposal for standardized grading of chronic changes in native kidney biopsy specimens. Kidney International. 2017/04/01/ 2017;91(4):787–789. 10.1016/j.kint.2017.01.002

15. Ingegnoli F, Ardoino I, Boracchi P, et al. Nailfold capillaroscopy in systemic sclerosis: data from the EULAR scleroderma trials and research (EUSTAR) database. Microvasc Res. Sep 2013;89:122–8. doi:10.1016/j.mvr.2013.06.003

16. Antonios TF, Nama V, Wang D, et al. Microvascular remodelling in preeclampsia: quantifying capillary rarefaction accurately and independently predicts preeclampsia. Am J Hypertens. Sep 2013;26(9):1162–9. doi:10.1093/ajh/hpt087

17. Stewart J, Kohen A, Brouder D, et al. Noninvasive interrogation of microvasculature for signs of endothelial dysfunction in patients with chronic renal failure. American Journal of Physiology-Heart and Circulatory Physiology. 2004;287(6):H2687–H2696. doi:10.1152/ajpheart.00287.2004

18. Li W, Schram MT, Berendschot TTJM, et al. Type 2 diabetes and HbA1c are independently associated with wider retinal arterioles: the Maastricht study. Diabetologia. 2020/07/01 2020;63(7):1408–1417. doi:10.1007/s00125-020-05146-z

19. Houben AJHM, Schaper NC, Slaaf DW, et al. Skin blood cell flux in insulin-dependent diabetic subjects in relation to retinopathy or incipient nephropathy. European Journal of Clinical Investigation. 1992;22(1):67–72. 10.1111/j.1365-2362.1992.tb01938.x

20. Pober JS, Sessa WC. Inflammation and the blood microvascular system. Cold Spring Harb Perspect Biol. Oct 23 2014;7(1):a016345. doi:10.1101/cshperspect.a016345

21. Six I, Flissi N, Lenglet G, et al. Uremic Toxins and Vascular Dysfunction. Toxins. 2020;12(6):404.

22. Martens RJH, Houben A, Kooman JP, et al. Microvascular endothelial dysfunction is associated with albuminuria: the Maastricht Study. J Hypertens. May 2018;36(5):1178–1187. doi:10.1097/hjh.0000000000001674

23. Seliger SL, Salimi S, Pierre V, et al. Microvascular endothelial dysfunction is associated with albuminuria and CKD in older adults. BMC Nephrol. Jul 13 2016;17(1):82. doi:10.1186/s12882-016-0303-x

24. Uemura O. Detection of haematuria and proteinuria reflects injury to significantly different glomerular surface areas: a quantitative hypothesis. Clin Kidney J. Aug 2025;18(8):sfaf245. doi:10.1093/ckj/sfaf245

25. Baaten C, Vondenhoff S, Noels H. Endothelial Cell Dysfunction and Increased Cardiovascular Risk in Patients With Chronic Kidney Disease. Circ Res. Apr 14 2023;132(8):970–992. doi:10.1161/circresaha.123.321752

26. Martens CR, Edwards DG. Peripheral vascular dysfunction in chronic kidney disease. Cardiol Res Pract. 2011;2011:267257. doi:10.4061/2011/267257

27. Theodorakopoulou MP, Iatridi F, Stavropoulos K, et al. Structural and Functional Capillary Integrity, Arterial Stiffness and Central Hemodynamics in CKD Patients With and Without Nocturnal Hypertension. American Journal of Hypertension. 2025;38(8):580–587. doi:10.1093/ajh/hpaf043

28. Li W, Schram MT, Sörensen BM, et al. Microvascular Phenotyping in the Maastricht Study: Design and Main Findings, 2010-2018. Am J Epidemiol. Sep 1 2020;189(9):873–884. doi:10.1093/aje/kwaa023

29. Choi PJ, Brunt VE, Fujii N, et al. New approach to measure cutaneous microvascular function: an improved test of NO-mediated vasodilation by thermal hyperemia. Journal of Applied Physiology. 2014;117(3):277–283. doi:10.1152/japplphysiol.01397.2013

